# Effect of losartan on hospitalized patients with COVID-19-induced lung injury: A randomized clinical trial

**DOI:** 10.1101/2021.08.25.21262623

**Authors:** Michael A. Puskarich, Nicholas E. Ingraham, Lisa H Merck, Brian E Driver, David A. Wacker, Lauren Page Black, Alan E. Jones, Courtney V. Fletcher, Andrew M. South, Andrew C. Nelson, Thomas A Murray, Christopher J. Tignanelli, Christopher Lewandowski, Joseph Farhat, Justin L. Benoit, Dana Byrne, Alex Hall, Ronald A. Reilkoff, Michelle H. Biros, Kartik Cherabuddi, Jeffrey G. Chipman, Timothy W. Schacker, Tyler Bold, Kenneth Beckman, Ryan Langlois, Matthew T. Aliota, Faheem W. Guirgis, James Galbriath, Margaret Beyer, Chas Salmen, Brian Roberts, David Wright, Helen T. Voelker, Joseph S. Koopmeiners

## Abstract

**Background:** SARS-CoV-2 viral entry may disrupt angiotensin II (Ang II) homeostasis in part via ACE2 downregulation, potentially contributing to COVID-19 induced lung injury. Preclinical models of viral pneumonias that utilize ACE2 demonstrate Ang II type 1 receptor (AT1R) blockade mitigates lung injury, though observational COVID-19 data addressing the effect of AT1R blockade remain mixed.

**Methods:** Multicenter, blinded, placebo-controlled randomized trial of losartan (50 mg PO twice daily for 10 days) versus placebo. Hospitalized patients with COVID-19 and a respiratory sequential organ failure assessment score of at least 1 and not already taking a renin-angiotensin-aldosterone system (RAAS) inhibitor were eligible. The primary outcome was the imputed partial pressure of oxygen to fraction of inspired oxygen (PaO_2_/FiO_2_) ratio at 7 days. Secondary outcomes included ordinal COVID-19 severity, oxygen, ventilator, and vasopressor-free days, and mortality. Losartan pharmacokinetics (PK) and RAAS components [Ang II, angiotensin-(1–7) (Ang-(1–7)), ACE, ACE2] were measured in a subgroup of participants.

**Findings:** From April 2020 - February 2021, 205 participants were randomized, 101 to losartan and 104 to placebo. Compared to placebo, losartan did not significantly affect PaO_2_/FiO_2_ ratio at 7 days [difference of -24.8 (95% -55.6 to 6.1; p=0.12)]. Losartan did not improve any secondary clinical outcome, but worsened vasopressor-free days. PK data were consistent with appropriate steady-state concentrations, but we observed no significant effect of losartan on RAAS components.

**Interpretation:** Initiation of orally administered losartan to hospitalized patients with COVID-19 and acute lung injury does not improve PaO_2_ / FiO_2_ ratio at 7 days. These data may have implications for ongoing clinical trials.

**Trial Registration:** Losartan for Patients With COVID-19 Requiring Hospitalization (NCT04312009), https://clinicaltrials.gov/ct2/show/NCT04312009

## Background

SARS-CoV-2 has infected over 195 million persons, causing over 4 million deaths.^1^ SARS-CoV-2 enters respiratory epithelial cells via angiotensin-converting enzyme 2 (ACE2), a major component of the renin-angiotensin-aldosterone system (RAAS).^2,3^ ACE2 is responsible for degradation of angiotensin II (Ang II), a proinflammatory vasoconstrictor, into angiotensin-(1– 7) [Ang-(1–7)] and angiotensin-(1–9), which generally oppose Ang II’s effects.^2,4^ Early data suggested Ang II levels may be associated with viral load and degree of lung injury in COVID-19 patients,^4^ though more recent and methodologically robust data demonstrate mixed findings.^5–7^

Preclinical models of viral pneumonias affecting ACE2, including severe acute respiratory syndrome coronavirus (SARS-CoV), demonstrate Ang II type 1 receptor (AT1R) blockade reduces lung injury and death.^8–10^ Observational studies of antecedent angiotensin-converting enzyme inhibitor (ACE-i) or AT1R blocker (ARB) use and disease severity and mortality in COVID-19, meanwhile, demonstrate mixed results.^11–17^ RAAS inhibition in COVID-19, therefore, remains an ongoing area of controversy.

We hypothesized losartan treatment might reduce lung injury and improve clinical outcomes in hospitalized patients with COVID-19 by restoring Ang II and Ang-(1–7) homeostasis. To test this hypothesis, we conducted a multicenter blinded randomized placebo-controlled clinical trial of patients hospitalized with COVID-19 not already taking a RAAS inhibitor. The primary objective was to test if losartan improves the PaO_2_/FiO_2_ (P/F) ratio at 7 days. We also sought to determine if losartan affects biochemical markers (including RAAS components), severity of illness, and mortality.

## Methods

### Study design

We conducted a prospective multicenter blinded randomized placebo-controlled trial of hospitalized patients with COVID-19 at 13 hospitals in the United States from April 2020 to February 2021 following CONSORT guidelines.^18^ The protocol was approved by a central institutional review board (Advarra, Pro00042757) and conducted following good clinical practice guidelines under the oversight of an independent data safety monitoring board. All participants or their legally authorized representatives (LARs) provided written electronic informed consent. The trial was conducted under the authority of the Food and Drug Administration (IND 148365) and registered on clinicaltrials.gov prior to initialization (NCT043112009).

### Participants

Consecutive patients presenting to a participating institution with a positive clinical RT-PCR SARS-CoV-2 result were assessed for eligibility. Participants >=18 years old were potentially eligible if exhibiting at least one CDC-recognized symptom of COVID-19.^19^ Exclusion criteria included: active outpatient treatment with an ACE-i, ARB, or aliskiren; prior adverse reaction to ACE-i/ARB; pregnancy, breastfeeding, or lack of contraception; history of dialysis, stage IV chronic kidney disease, or estimated glomerular filtration rate (eGFR) <30 mL/min/1.73 m^2^; potassium >5.0 mmol/L; history of severe liver disease; enrollment in another blinded clinical trial for COVID-19; lack of informed consent; inability to randomize within 48 hours of admission or positive test; oxygen saturation at baseline; or respiratory sequential organ failure assessment (SOFA) score of <1 (defined as P/F of <400, utilizing SaO2 if PaO2 is unavailable).^20,21^

### Screening and consent

Electronic health records (EHR) were screened by manual chart review by trained research personnel. Due to limitations in personal protective equipment, informed consent was typically conducted remotely via telephone or video teleconference, and documentation was maintained using a 21 CFR part 11 compliant electronic consent (eConsent) platform (REDCap).^22^

### Randomization and blinding

Enrolled participants were randomized using permuted blocks of randomly varying sizes (2, 4, or 6) stratified by site and age (>=60 or <60 years). A central randomization website generated treatment allocations in a 1:1 ratio. All participants and study personnel were blinded except statisticians preparing the randomization and investigational pharmacists.

### Intervention

The intervention was losartan 50 mg PO twice daily (100 mg daily total) versus equally appearing placebo, yielding an expected 70% inhibition of AT1R.^23^ Study drug was shipped to sites as pills and prepared locally by an unblinded pharmacist in suspension per the manufacturer’s package insert. Study drug was administered for 10 days if eGFR >60 mL/min/1.73 m^2^, once daily for eGFR 30–60, and discontinued if eGFR decreased below 30, following discharge, or by a blinded investigator if a drug-related serious adverse effect (SAE) was suspected.

### Primary efficacy outcome

The primary outcome was the PaO_2_/FiO_2_ ratio on day 7. If lacking, PaO_2_ was imputed using the method of Pandharipande (for positive pressure ventilation) or Gadrey (no positive pressure).^21,24^ Participants discharged prior to day 7 were provided a home pulse oximeter and contacted via phone.

### Secondary outcomes

Oxygen, ventilator, and vasopressor-free days (to 10 days), time to discharge, 7-point ordinal scale of COVID-19 severity,^25^ and 28-day mortality were measured. A subset of participants (limited by local biohazard capacity) underwent biospecimen collection.

### Safety monitoring

Creatinine and potassium were measured on days 0, 1, 3, and 7. Acute kidney injury was defined as an increase in serum creatinine of 0.3 mg/dL or 1.5-times baseline.^26^ Blood pressures and medical charts were reviewed daily for up to 15 days post-randomization.

### Pharmacokinetic measurements

Blood samples were obtained at 2, 4, and 6 hours after a dose of losartan or placebo (50 mg) in a subgroup blinded to treatment allocation. Plasma concentrations of losartan and its active carboxy metabolite, carboxylosartan (LCA) were quantified by a validated LC-MS/MS assay with a lower limit of quantification of 3 ng/mL at the University of Nebraska Medical Center.

### RAAS measurements

Blood was collected in an EDTA tube containing a cocktail of protease inhibitors validated with the intended assays and red-top tubes per best practices.^27–29^ Extracted plasma and serum were stored at -80°C and sent to the CLIA-certified Biomarker Analytical Core at Atrium Health Wake Forest Baptist for analysis. The plasma was thawed on ice, extracted on Sep-Pak C18 columns (Waters Corp., Milford, Massachusetts, USA), and the eluted fractions analyzed with radioimmunoassays.^28^ Plasma Ang II was measured using the Immuno-Biological Lab, Inc. kit (IBL America, Minneapolis, MN 55432), while plasma Ang-(1–7) was measured using an antibody produced by the laboratory and validated with LC-MS/MS. Serum ACE and ACE2 activity were analyzed using established methods.^27,28^

### Viral load

Consistent with our prior work, we report normalized Ct value for SARS-CoV-2 targets compared to human RP internal sample control, providing a relative value to compensate for sampling and extraction quality.^30^

### Statistical analysis

Statistical analyses were performed using SAS (version 9.4 or newer) or R (version 4.0.3 or newer).^31^ All statistical tests were two-tailed with p-values less than 0.05 considered statistically significant. Baseline clinical characteristics are summarized using descriptive statistics. All analyses were conducted using intent-to-treat principles. We tested the effect of losartan on P/F ratio at Day 7 using predictive modeling, imputing zero for participants that died early to penalize for death. Missing values in alive participants were multiply imputed 20 times using predictive mean matching. The predictive models included study arm, location of enrollment, baseline hypertension, assigned sex, age, body mass index, baseline P/F ratio, and days from randomization to discharge. Secondary unadjusted analysis of the primary outcome was carried out using Welch’s t-test.

Longitudinal secondary endpoints were analyzed using generalized linear mixed models or generalized estimating equations, adjusting for corresponding baseline measurements. Mortality was summarized using Kaplan-Meier plots and compared using the log-rank test and unadjusted Cox models to estimate hazard ratios. Time to hospital discharge, with death in hospital or discharge to comfort care as a competing risk, was evaluated using cumulative incidence plots and Fine-Gray’s test (rho = 0), the competing risks’ analog to the log-rank test. Ordinal outcomes were analyzed using proportional odds regression.

Pharmacokinetic characteristics were determined using the trapezoidal rule and linear regression (Phoenix WinNonLin v8.3, Certara, Princeton, NJ, USA). RAAS data were analyzed by jointly modeling RAAS components and time-to-discharge using the ‘JM’ package in R on the natural-log scale using linear mixed effects models with terms for treatment assignment, baseline values, and a linear term for day. Viral load was analyzed using linear mixed models with a visit-by-treatment group interaction and a global Wald test for overall treatment statistical significance.

### Power and sample size

Due to a paucity of data at study design, we based expected variance of P/F on prior studies of viral-induced acute lung injury, considering standard deviations (SD) between 50-125.^32–35^ At a sample size of 200 with a 1:1 allocation ratio, we calculated 90% power to detect a difference of 70 in the P/F ratio assuming SD 150, and 80% to detect a 50-unit difference, assuming SD 125.

## Results

Of the 4,113 patients screened, 3,672 met exclusion criteria (**Figure 1**). Of those eligible, 47% (208) provided consent to participate. Three developed post-enrollment exclusions, leaving 205 randomized participants (104 placebo and 101 losartan), representing the intent-to-treat cohort. All participants were followed until completion or withdrawal.

**Figure 1.**
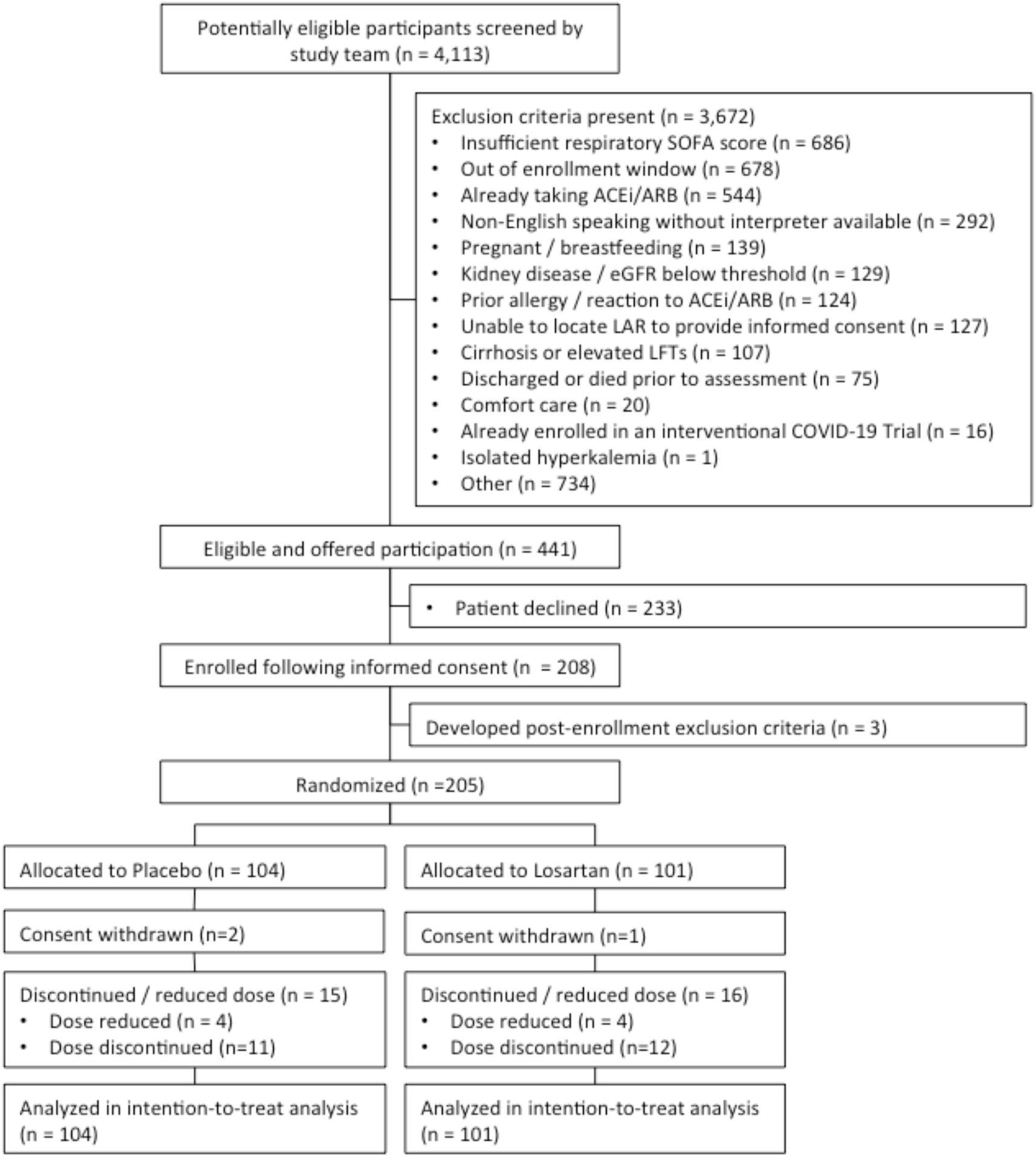
Study flow diagram of patient enrollments and randomization

Demographics and clinical characteristics were fairly well matched (**Table 1**), though more patients in the losartan arm were enrolled in the ICU (16.8% vs 8.7%). Participants were middle aged (mean age 55 years) and racially and ethnically diverse (33% Black and 21% Hispanic). A high proportion were overweight (89%) or obese (63%), while almost half carried a preexisting diagnosis of hypertension (40%).

**Table 1:** Participant Demographic and Comorbid Condition Summaries. Categorical summaries reflect the number (%) {# missing} and continuous summaries reflect mean (standard deviation) {# missing}

|  |  |  |
| --- | --- | --- |
|  | *Placebo* | *Losartan* |
| Number Enrolled | 104 | 101 |
| Location of Enrollment |  |  |
| ED | 20 (19.2%) | 20 (19.8%) |
| Floor | 63 (60.6%) | 59 (58.4%) |
| Step-Down or Intermediate | 12 (11.5%) | 5 (5%) |
| ICU | 9 (8.7%) | 17 (16.8%) |
| Demographic Summary |  |  |
| Female | 41 (39.4%) {0} | 41 (40.6%) {0} |
| Age (years) | 56.4 (15) {0} | 53.8 (16.1) {0} |
| 18-34 | 5 (4.8%) | 13 (12.9%) |
| 35-54 | 41 (39.4%) | 42 (41.6%) |
| 55-64 | 34 (32.7%) | 23 (22.8%) |
| 65-89 | 20 (19.2%) | 22 (21.8%) |
| 90+ | 4 (3.8%) | 1 (1%) |
| BMI (kg/m<sup>2</sup>) | 33.1 (8.9) {1} | 34 (9.9) {0} |
| Underweight (<18.5) | 1 (1%) | 0 (0%) |
| Normal weight (18.5 - <25) | 12 (11.5%) | 10 (9.9%) |
| Overweight (25 - <30) | 26 (25%) | 26 (25.7%) |
| Class I Obesity (30 - <35) | 36 (34.6%) | 30 (29.7%) |
| Class II Obesity (35 - <40) | 7 (6.7%) | 16 (15.8%) |
| Class III Obesity (40+) | 21 (20.2%) | 19 (18.8%) |
| Race |  |  |
| White | 47 (45.2%) | 35 (34.7%) |
| Black or African American | 30 (28.8%) | 37 (36.6%) |
| Native American or Alaskan Native | 1 (1%) | 0 (0%) |
| Asian | 2 (1.9%) | 7 (6.9%) |
| Hispanic | 18 (17.3%) | 19 (18.8%) |
| Other/Unknown | 6 (5.8%) | 3 (3%) |
| Ethnicity |  |  |
| Non-Hispanic or Latino | 76 (73.1%) | 80 (79.2%) |
| Hispanic or Latino | 23 (22.1%) | 20 (19.8%) |
| Unknown | 5 (4.8%) | 1 (1%) |
| Insured | 80 (76.9%) {1} | 83 (82.2%) {1} |
| Medicaid | 18 (17.3%) {0} | 18 (17.8%) {0} |
| Medicare | 32 (30.8%) {0} | 21 (20.8%) {0} |
| Private | 42 (40.4%) {0} | 59 (58.4%) {0} |
| Comorbid Condition Summary |  |  |
| Coronary Artery Disease | 7 (6.7%) {0} | 9 (8.9%) {0} |
| Hypertension | 46 (44.2%) {0} | 37 (36.6%) {0} |
| On HTN Medication | 29 (27.9%) {58} | 21 (20.8%) {66} |
| Congestive Heart Failure | 4 (3.8%) {0} | 5 (5%) {0} |
| Pulmonary Hypertension | 6 (5.8%) {0} | 4 (4%) {0} |
| Asthma | 14 (13.5%) {0} | 13 (12.9%) {0} |
| Chronic Obstructive Pulmonary Disease | 10 (9.6%) {0} | 14 (13.9%) {0} |
| Chronic Bronchitis | 2 (1.9%) {0} | 6 (5.9%) {0} |
| Obstructive Sleep Apnea | 22 (21.2%) {0} | 12 (11.9%) {0} |
| Diabetes Mellitus | 26 (25%) {0} | 21 (20.8%) {1} |
| Tobacco or Nicotine User | 9 (8.7%) {0} | 4 (4%) {0} |

Primary outcome data were missing in 15.4% of the placebo arm and 9.9% of the losartan arm, due to an inability to contact the participant on day 7 or participant failure to record pulse oximetry values. This difference reflects a higher proportion of participants discharged prior to day 7 in the placebo arm (59.6% vs. 48.5%). Three participants (2.9%) in the placebo arm and 2 (2.0%) in the intervention arm died prior to day 7.

Based on raw data, P/F ratio did not differ between the placebo and intervention arms [297 (95% CI 196-366) vs 297 (95% CI 130-366)] (**Figure 2, Supplementary Table 1**). In the generalized model representing our primary analysis, the estimated effect of losartan on the P/F ratio was -24.8 (95% CI -55.6 to 6.1; p=0.12), with positive values indicating improvement. Secondary complete analyses affected neither the direction, magnitude, or the statistical significance of the estimate.

**Figure 2.**
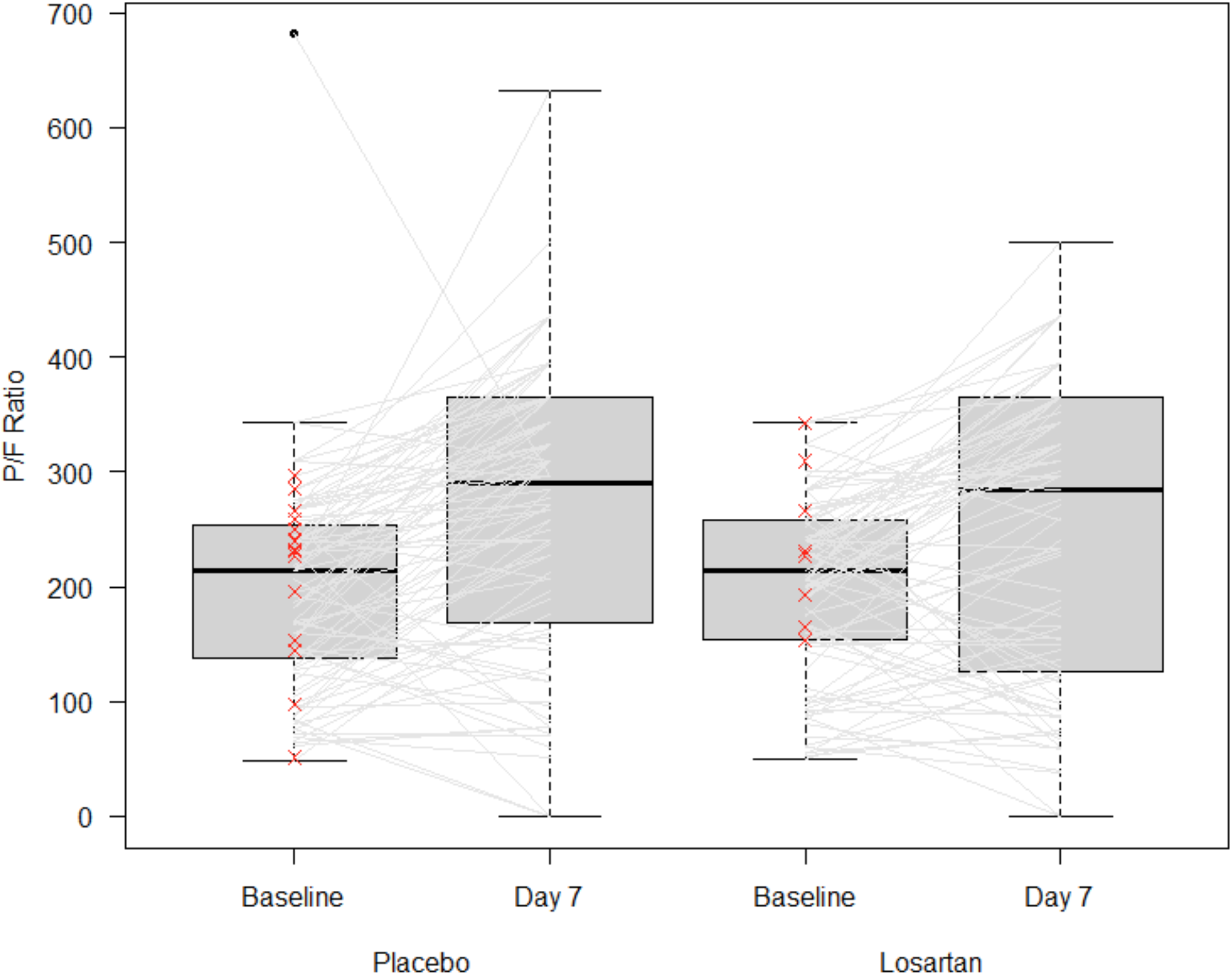
Change in P/F ratios from day 0 to 7, by treatment allocation. Participants with missing day 7 value are indicated with a red “x” at their baseline value, otherwise each participant’s baseline and day 7 values are connected with a grey line.

We observed no difference in time to hospital discharge or in-hospital mortality (**Figure 3**). By day 28, 11 participants in the intervention arm died vs 9 in the placebo arm, equilibrating to 11 in each group by day 90. There were no differences in freedom from oxygen or mechanical ventilation in the first 10 days following randomization, and the ordinal outcome did not differ between treatment groups (data not shown).

**Figure 3.**
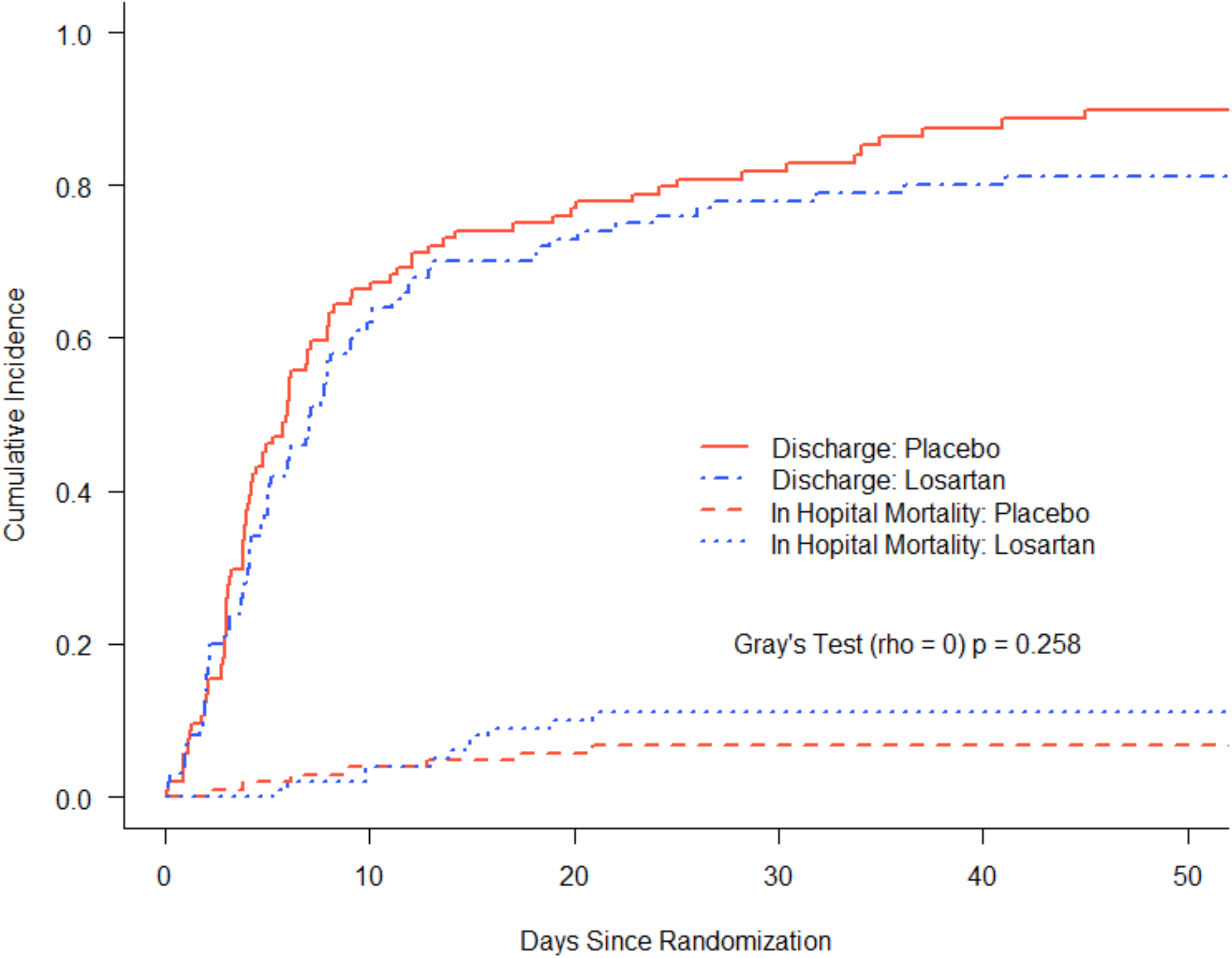
Time-to-event curves of hospital discharge and survival, comparing placebo (red lines) and losartan (blue lines). No statistically differences between groups were observed (p=0.258)

More patients in the losartan arm required vasopressors (20 versus 11, p = 0.08), contributing to fewer vasopressor-free days in the intervention group (8.7 versus 9.4, p = 0.04, **Table 2**). Adverse events did not differ significantly by treatment allocation (**Supplementary Table 2**), though a numerically higher number of cardiovascular SAEs were noted in the losartan arm. Acute kidney injury by KDIGO was more common in the interventional arm (19.4% versus 8.8%, p=0.04).

**Table 2:**
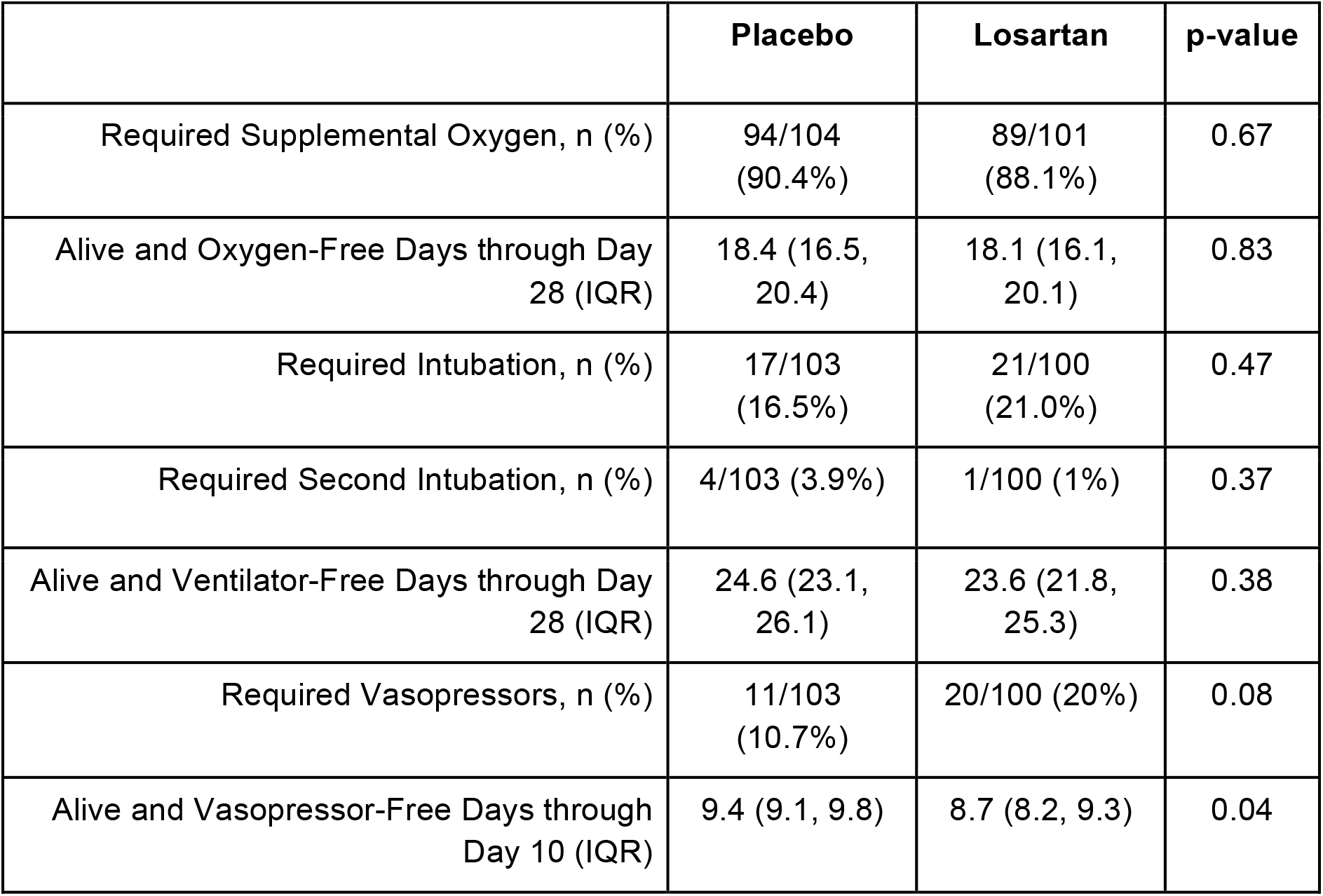
Alive and Intervention-Free Days Analyses

| **Table 2: Alive and Intervention-Free Days Analyses** |  |  |  |
| --- | --- | --- | --- |
|  | **Placebo** | **Losartan** | **p-value** |
| Required Supplemental Oxygen, n (%) | 94/104<br>(90.4%) | 89/101<br>(88.1%) | 0.67 |
| Alive and Oxygen-Free Days through Day<br>28 (IQR) | 18.4 (16.5,<br>20.4) | 18.1 (16.1,<br>20.1) | 0.83 |
| Required Intubation, n (%) | 17/103<br>(16.5%) | 21/100<br>(21.0%) | 0.47 |
| Required Second Intubation, n (%) | 4/103 (3.9%) | 1/100 (1%) | 0.37 |
| Alive and Ventilator-Free Days through Day<br>28 (IQR) | 24.6 (23.1,<br>26.1) | 23.6 (21.8,<br>25.3) | 0.38 |
| Required Vasopressors, n (%) | 11/103<br>(10.7%) | 20/100 (20%) | 0.08 |
| Alive and Vasopressor-Free Days through<br>Day 10 (IQR) | 9.4 (9.1, 9.8) | 8.7 (8.2, 9.3) | 0.04 |

Viral load did not differ by treatment (**Supplementary Figure 1**). We detected no losartan or LCA in any placebo treated participant (n=3), while losartan and LCA pharmacokinetics were consistent with expected values in those treated with losartan (n=7; **Supplementary Figure 2**, and **Supplementary Table 3**). Analysis of RAAS biomarkers demonstrated no significant effect of losartan on the change in Ang-(1–7) (p=0.953), Ang II (p=0.715) (p=0.386), or ACE2 (p=0.538) over time(**Supplementary Table 4**).

## Discussion

In this multicenter, blinded, randomized, placebo-controlled clinical trial, 205 hospitalized participants with COVID-19 and acute lung injury not already taking RAAS inhibitors were randomized to oral losartan at its maximal FDA-approved dose to test the hypothesis that AT1R blockade improves pulmonary function. Despite a moderate to severely ill cohort, well matched P/F ratios, and pharmacologically relevant steady-state concentrations, we found no evidence that treatment with losartan improves lung injury or clinical outcomes, nor significantly affects major circulating RAAS components. We recently published our experience in an analogous trial in the outpatient setting.^30^ Though this trial was terminated early, losartan did not reduce hospital admissions, dyspnea, nor quality of life. Given a relatively healthy cohort, a low prevalence of hypertension, and a low event rate, it remained unclear whether more severely ill patients with hypothetically greater Ang II expression relative to Ang-(1–7) might benefit from losartan. This trial advances those findings. Specifically, our results do not support the hypothesis that losartan effectively mitigates viral-induced acute lung injury in COVID-19, with uncertain implications for other, potentially more potent, agents that target the RAAS.

It is important to note that we observed two potentially harmful effects of losartan on hemodynamics and kidney function. However, a higher proportion of participants in the intervention arm were enrolled in the ICU, indicating potential small imbalances in severity of illness at enrollment. Further, while the losartan group was more likely to meet acute kidney injury criteria by KDIGO, this finding may indicate expected physiologic changes in intraglomerular blood flow rather than actual tissue injury per se. Nevertheless, in the absence of benefit and potential for harm, we conclude losartan treatment is not indicated in this setting.

In light of our results, it is important to consider the pharmacology and biochemical effects of the intervention. The observed pharmacokinetics of losartan and LCA are consistent with reported values^23^ and maximal AT1R blockade.^39^ As the half-lives of losartan and LCA are ∼2 and 6 hours, and time to maximal concentrations were 1 and 4 hours, steady-state was achieved in the first day of dosing. Despite this, plasma Ang II and Ang-(1–7) concentrations and serum ACE and ACE2 activities were not affected. This may be interpreted that the intervention window was simply too narrow to observe the hypothesized biochemical effects. Alternative explanations include poor specificity of circulatory RAAS (unknown tissue source) or relative lack of RAAS dysregulation at baseline in this population.^4^ More recent observations may suggest SARS-CoV-2 does not induce unique, clinically relevant RAAS alterations, even in those who develop severe disease. Future analyses could investigate whether the circulatory RAAS is a mediator for treatment-induced effects on the outcomes.^40^

These results stand in contrast to an open-label trial of telmisartan that demonstrated significantly reduced 30-day mortality in the treatment arm (4.3% vs 22.5%).^31,36^ However, that study was powered to detect a reduction in C-reactive protein, was unblinded, excluded ICU patients, and was terminated early due to slow recruitment, limiting the interpretation of the observed mortality reduction. Two additional trials, meanwhile, examined discontinuation versus continuation of RAAS inhibitors on admission for COVID-19, and found no significant differences between groups, further supporting current guidelines against routine discontinuation of these medications.^37,38^ Our study complements and advances these findings.

There are several limitations to consider with this study. The trial was initiated early in the pandemic, and temporal changes in clinical care including introduction of dexamethasone and remdesivir may have biased the trial towards the null. Our findings may also be consistent with the hypothesis that the RAAS does not play a significant role in SARS-CoV-2-related acute lung injury relative to other inflammatory pathways. Our choice of primary efficacy outcome required differential imputation methods based on whether or not the participant was treated with positive pressure ventilation, potentially affecting the results. However, the lack of effect on oxygen or mechanical ventilation-free days decreases the likelihood a larger study would identify clinically meaningful effects. While we detected sufficient concentrations of losartan, it remains possible lung parenchymal penetration is limited, mitigating potential efficacy. The relatively small sub-group undergoing RAAS measurement may have been underpowered to detect treatment effects. It remains possible that certain subgroups may benefit from ARB treatment that we could not identify due to inadequate subgroup sample size, including patients who may have RAAS dysregulation at baseline prior to infection. Finally, we cannot necessarily generalize these findings to other ARBs, ACE inhibitors, or other agents that modulate the RAAS.

## Conclusion

Initiation of oral losartan to hospitalized patients with COVID-19 and acute lung injury does not improve PaO_2_ / FiO_2_ ratio at 7 days. These data have implications for ongoing clinical trials.

## Data Availability

Deidentified data will be fully available upon peer reviewed publication at the Data Repository for the University of Minnesota

## Acknowledgements

Lee Winchester and Timothy Mykris from the Antiviral Pharmacology Laboratory, University of Nebraska Medical Center for quantitation of losartan and carboxylosartan concentrations, and Dr. TanYa Gwathmey, Dr. Bridget Brosnihan, and Pam Dean from the Biomarker Analytical Core at Atrium Health Wake Forest Baptist for analysis of RAAS components. We would like to acknowledge the project manager Dave Ankarlo.

---

**Supplementary Table 1:** PaO_2_/FiO_2_ ratio at Baseline with Day 7 results. Reported quantities reflect Median (IQR) or N (%).

|  | **Placebo**<br>(n = 104) | **Losartan**<br>(n = 101) |
| --- | --- | --- |
| P/F Ratio at Baseline | 214.1 [138.4 - 253.4] | 214.1 [153.6 - 258.4] |
| Discharged on or before Day 7 | 62 (59.6%) | 49 (48.5%) |
| Hospitalized through Day 7 | 39 (37.5%) | 50 (49.5%) |
| Dead on or before Day 7 | 3 (2.9%) | 2 (2.0%) |
| P/F Ratio Missing on Day 7 (Alive) | 16 (15.4%) | 10 (9.9%) |
| P/F Ratio on Day 7 (Alive and Non-Missing) | 296.5 [196.0 - 365.7] | 296.5 [130.0 - 365.7] |

**Supplementary Table 2.** Inpatient Non-Fatal Adverse Events per person-day, day 1-15

|  | Losartan |  | Placebo |  |  |
| --- | --- | --- | --- | --- | --- |
|  | Number of Events | Rate (SD) | Number of Events | Rate (SD) | P-value |
| **Adverse events (including serious adverse events)** |  |  |  |  |  |
| Cardiovascular | 26 | 0.021 (0.053) | 24 | 0.021 (0.077) | 0.96 |
| Endocrine | 2 | 0.001 (0.010) | 1 | 0.001 (0.007) | 0.51 |
| Ear, Nose, and Throat | 0 | 0.000 (0.000) | 1 | 0.001 (0.007) | 0.32 |
| Gastrointestinal | 10 | 0.007 (0.021) | 3 | 0.002 (0.011) | 0.04 |
| Genitourinary | 3 | 0.002 (0.011) | 1 | 0.002 (0.016) | 0.84 |
| Hematologic | 9 | 0.006 (0.022) | 6 | 0.005 (0.028) | 0.72 |
| Musculoskeletal | 0 | 0.000 (0.000) | 0 | 0.000 (0.000) |  |
| Neurologic | 4 | 0.003 (0.013) | 4 | 0.003 (0.016) | 0.97 |
| Renal | 20 | 0.014 (0.051) | 21 | 0.017 (0.055) | 0.68 |
| Respiratory | 30 | 0.023 (0.051) | 27 | 0.023 (0.064) | 0.91 |
| Skin | 0 | 0.000 (0.000) | 2 | 0.001 (0.009) | 0.16 |
| Total | 104 | 0.077 (0.154) | 90 | 0.074 (0.204) | 0.89 |
| **Serious Adverse Events Only** |  |  |  |  |  |
| Cardiovascular | 14 | 0.011 (0.035) | 6 | 0.005 (0.023) | 0.16 |
| Endocrine | 0 | 0.000 (0.000) | 0 | 0.000 (0.000) |  |
| Ear, Nose, and Throat | 0 | 0.000 (0.000) | 0 | 0.000 (0.000) |  |
| Gastrointestinal | 1 | 0.001 (0.007) | 0 | 0.000 (0.000) | 0.32 |
| Genitourinary | 0 | 0.000 (0.000) | 0 | 0.000 (0.000) |  |
| Hematologic | 1 | 0.001 (0.007) | 0 | 0.000 (0.000) | 0.32 |
| Musculoskeletal | 0 | 0.000 (0.000) | 0 | 0.000 (0.000) |  |
| Neurologic | 2 | 0.001 (0.009) | 1 | 0.001 (0.007) | 0.55 |
| Renal | 1 | 0.001 (0.007) | 2 | 0.002 (0.013) | 0.47 |
| Respiratory | 19 | 0.016 (0.044) | 21 | 0.019 (0.063) | 0.75 |
| Skin | 0 | 0.000 (0.000) | 0 | 0.000 (0.000) |  |
| Total | 38 | 0.030 (0.069) | 30 | 0.026 (0.082) | 0.68 |

**Supplementary Table 3.**
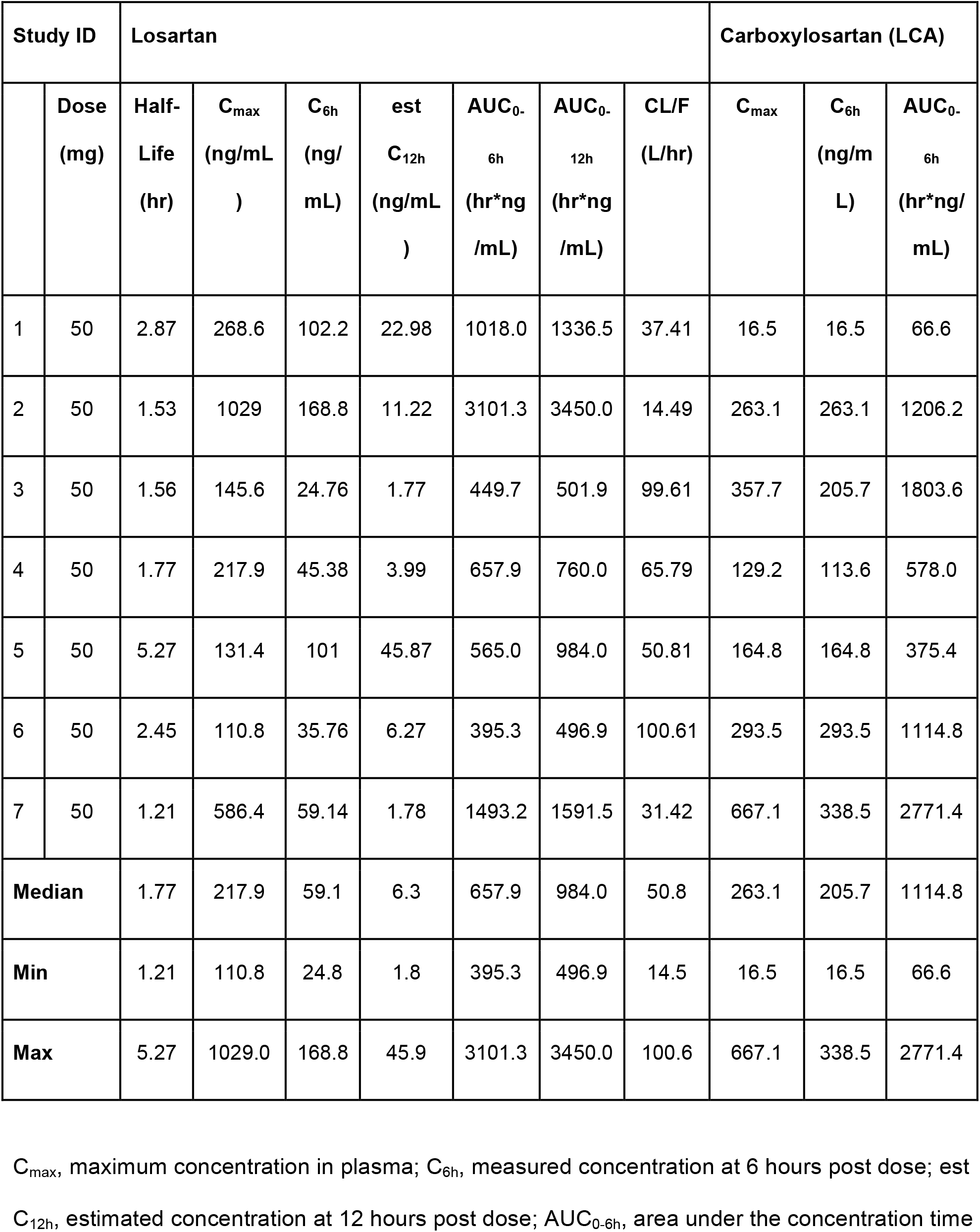
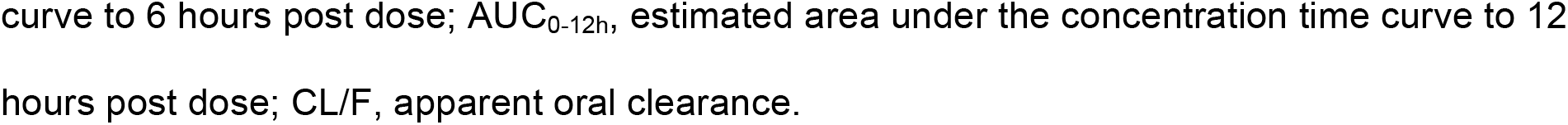
Losartan and carboxylosartan (LCA) pharmacokinetic characteristics

**Supplementary Table 4.** Association between treatment group and circulatory RAAS components after adjusting for baseline and time and jointly modeling the outcome and time-to-discharge (death treated as censored). Significant p values for log(baseline) indicate significant association of baseline measurements with later measurements, after controlling for treatment allocation and time.

| **Covariate** | **ACE** |  | **ACE2** |  | **Ang-(1–7)** |  | **Ang II** |  |
| --- | --- | --- | --- | --- | --- | --- | --- | --- |
|  | Ratio of Geometric Mean | p-value | Ratio of Geometric Mean | p-value | Ratio of Geometric Mean | p-value | Ratio of Geometric Mean | p-value |
| **Losartan** | 0.90<br>(0.72, 1.13) | 0.39 | 1.17<br>(0.71, 1.92) | 0.54 | 0.99<br>(0.63, 1.54) | 0.95 | 1.07<br>(0.76, 1.50) | 0.72 |
| **Log (baseline)** | 2.73<br>(2.37, 3.13) | <0.01 | 2.15<br>(1.87, 2.47) | <0.01 | 1.71<br>(1.44, 2.05) | <0.01 | 1.62<br>(1.23, 2.15) | <0.01 |
| **Day** | 1.02<br>(1.00, 1.04) | 0.12 | 1<br>(0.97, 1.04) | 0.98 | 0.98<br>(0.94, 1.01) | 0.16 | 1 (0.97, 1.03) | 0.96 |

ACE - Angiotensin-converting enzyme activity; ACE 2 - Angiotensin-converting enzyme 2 activity; Ang-(1–7) - angiotensin-(1–7) concentration; Ang II - angiotensin II concentration

**Supplementary Figure 1.**
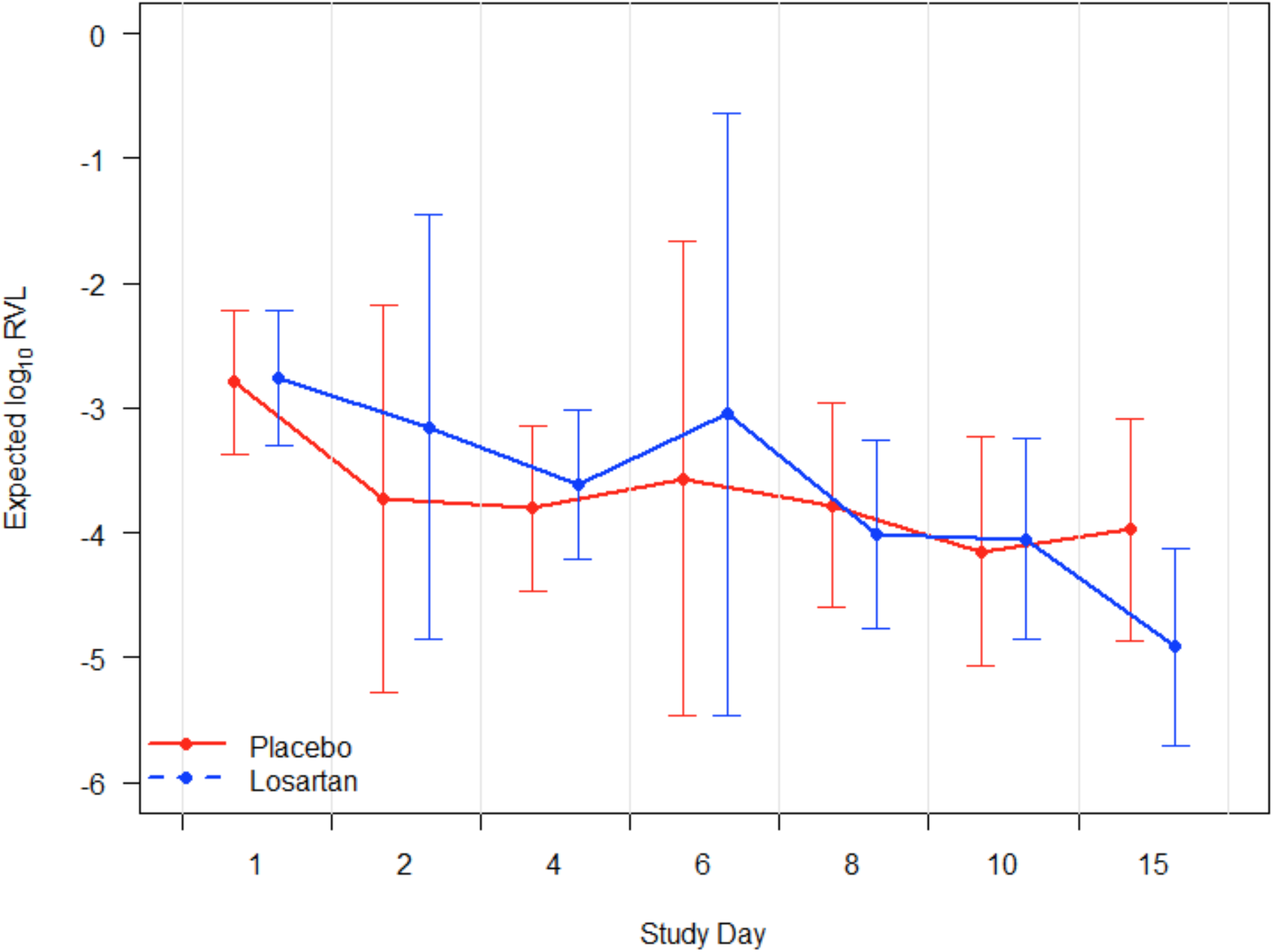
Effect of losartan on relative viral load (log10 scale); (relative viral load (RVL) is corrected to human marker DNA to control for specimen quality. Placebo is in red lines and losartan in blue lines with 95% CIs at each assessment. Losartan did not statistically significantly affect the cycle threshold or relative viral load overall or at any time point.

**Supplementary Figure 2.**
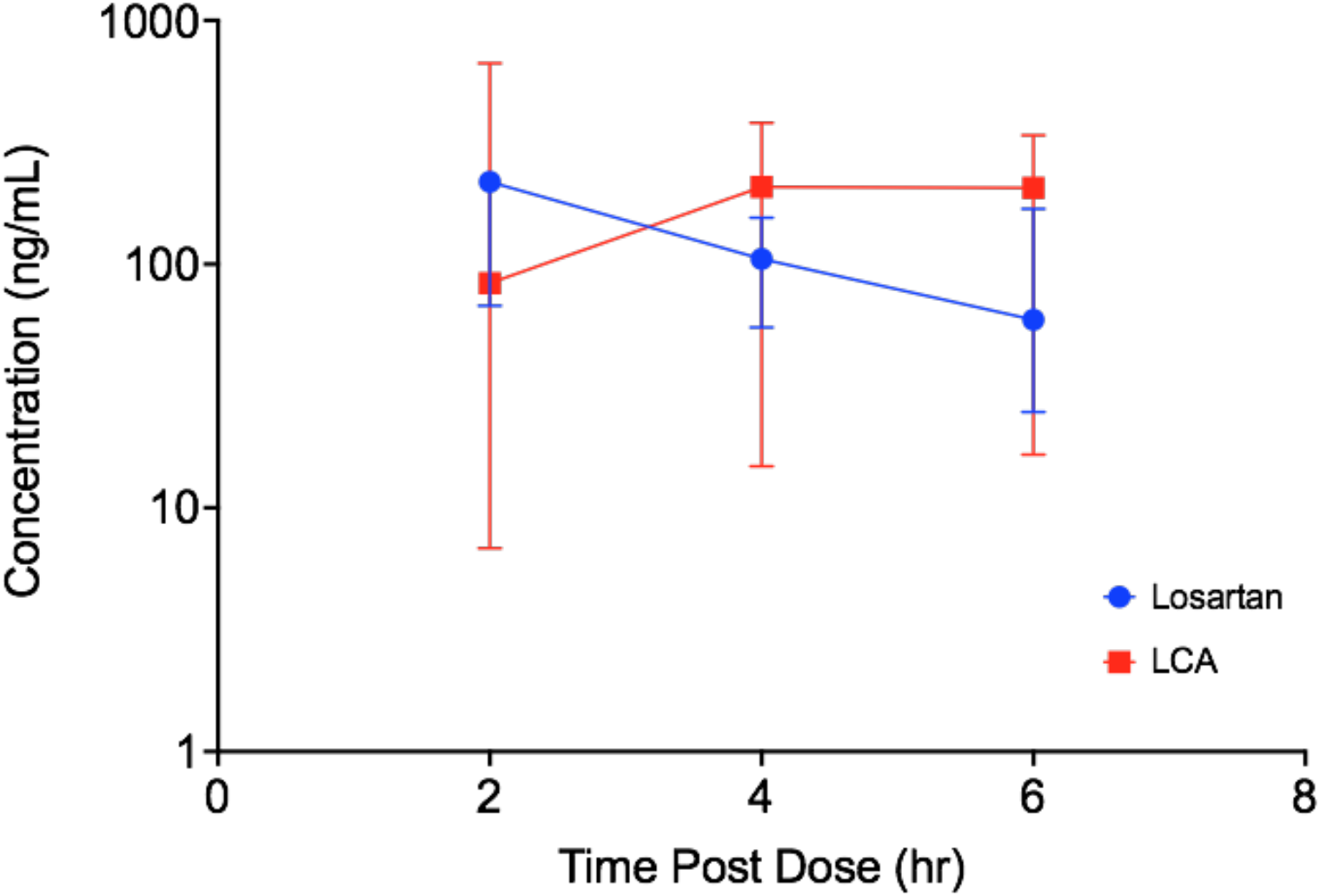
Losartan and carboxylosartan (LCA) concentrations (median and range).

